# COVID-19 risk variant associations with chromatin remodelling, DNA maintenance and surfactant genes are infection dependent in the lung

**DOI:** 10.1101/2023.05.03.23289478

**Authors:** Rachel K. Jaros, Evgeniia Golovina, Justin M. O’Sullivan

**Affiliations:** The Liggins Institute, The University of Auckland, Auckland 1023, New Zealand; Maurice Wilkins Centre for Molecular Biodiscovery, The University of Auckland, Auckland 1010, New Zealand; MRC Lifecourse Epidemiology Unit, University of Southampton, United Kingdom; Singapore Institute for Clinical Sciences, Agency for Science, Technology and Research (A*STAR), Singapore, Singapore; Australian Parkinson’s Mission, Garvan Institute of Medical Research, Sydney, New South Wales, Australia

**Author notes:** Corresponding senior author.

**Keywords:** SARS-CoV-2, COVID-19, regulatory genomics, GWAS, eQTL, chromatin, Hi-C

## Abstract

During viral infection the structure of host chromatin is modified. It is generally assumed that these chromatin modifications will affect variant-gene mapping, and therefore gene expression. What is not clear is how limitations imposed by host germline risk affect the expression changes that occur with infection induced chromatin remodelling. Critically, this lack of information extends to how germline variants associated with severe SARS-CoV-2 impact on tissue-specific gene expression changes in response to infection-induced chromatin conformation changes. Here we combined temporal chromatin conformation data from SARS-CoV-2 stimulated cells with a lung spatial-eQTL gene expression analysis to contextualise the functional effects and contributions of germline risk on the severe phenotypes observed in SARS-CoV-2. We identify changes in lung-specific SARS-CoV-2 risk variant-gene mapping across the infection time course. Our results provide evidence for infection-induced chromatin remodelling that impacts the regulation of genes associated with the severity of SARS-CoV-2 infection. The gene targets we identified are functionally involved in host chromatin modifications and maintenance and the expression of these genes is amplified by SARS-CoV-2-induced epigenetic remodelling. The effect of this remodelling includes transcriptional changes to gene targets such as *SMARCA4, NCOR1, DNMT1, DNMT3a, DAXX*, and *PIAS4*, all critical components of epigenetic control mechanisms and SARS-CoV-2 antiviral activity, along with several genes involved in surfactant metabolism. We show how severe-phenotype-associated eQTLs form and break in an infection time-course-dependent manner that mimics positive feedback loops connecting germline variation with the process of viral infection and replication. Our results provide a novel bridge between existing COVID-19 epigenetic research and demonstrate the critical role of epigenomics in understanding SARS-CoV-2-risk-associated gene regulation in the lung.

## Introduction

Genome-wide association studies of phenotypes observed in patients with COVID-19 have identified reproducible genetic associations indicating a genetic component to severity risk. Nonetheless, information about the functional effect of risk variants in the non-coding genome lacks two critical pieces of information, (1) the tissue-specificity of gene regulation and (2) the temporal effects of tissue-specific variant-gene pairs expressed under SARS-CoV-2 infected cells. Ninety-five gene targets associated with severity of infection were identified by COVID-19 GWAS‘s^1^. These were collectively mapped from 18 loci within the ‘hospitalisation‘ phenotype and 11 loci within the ‘severe‘ phenotype (i.e. hospitalised, respiratory support and/or death). Both act as a proxy for disease severity. The data, derived from the COVID-19 HGI^1-3^, is the most comprehensive GWAS on COVID-19 globally. The consortium used gene mapping techniques such as the closest gene, genes with a loss-of-function or missense variant in LD with a lead variant, genes with a fine-mapped cis-eQTL variant in GTEx^4^ and eQTL catalog that is in LD with a lead variant and highest gene prioritised by OpenTargetGenetics‘ V2G score^5^. None of these methods accounts for cell/tissue specificity relevant to the phenotype under study, nor do these studies consider epigenetic mechanisms that influence gene expression under SARS-CoV-2 conditions. These are arguably the most critical aspects of understanding the functional consequences of genetic risk^6^.

Mounting evidence^7-10^ suggests that epigenetic signals contribute to the severity of SARS-CoV-2 infection. Lung epithelial cells are the first target of SARS-CoV-2, and the A549^ACE2^ cell line constitutes a relevant cellular context for SARS-CoV-2 infection. Substantial chromatin condensation, increases in H3K9me3 and H3K27me3, and decreases in H3K9ac^9^ resulting in induction of inflammatory genes, and suppression of type 1 interferon-responsive antiviral genes and virus sensors was observed in SARS-CoV-2 infected A549^ACE2^ cells^7,9^. This is attributed to ORF8, which encodes a SARS-CoV-2 specific histone mimic^9^. In another infection-like model, macrophages treated with LPS and IFN-γ, genome-wide changes in chromatin loops, acetylation, and expression were observed by Hi-C over 24 hours at 7 time intervals^10^. Chromatin loops exhibit transcriptional control by increasing contact frequencies between enhancers and their target genes, which are partly regulated by external stimuli such as infection and substantial changes in gene transcription^10^. *SMARCA4*, identified as a SARS-CoV-2 risk gene target^6^ and by CRISPR screen as the second most proviral gene following ACE2^11^, forms part of the SWI/SNF remodelling complex that regulates chromatin accessibility and gene expression. Knockout of *SMARCA4* conferred resistance (i.e. proviral) to SARS-CoV-2 and was found to be unique to ACE2-receptor-mediated viruses^11^. Collectively, SARS-CoV-2 induces substantial chromatin remodelling by ORF8 and histone post-translational modifications and induces specific proviral gene programs. At the same time, host susceptibility in the germline could form a positive feedback loop in chromatin accessibility via gene targets such as *SMARCA4* and other chromatin remodellers.

Several functional assays applying whole-genome KO CRISPR screens, or specific regions^12^, for identifying SARS-CoV-2 regulators have been reported^11,13-18^. These screens varied in the viral isolate, CRISPR library and cell type. A meta-analysis showed a high level of cell-type specificity of the identified hits^13^. Regardless, these experiments uncovered critical host factors for the virus. Top proviral genes include *ACE2* and *CTSL* across all screens^11,13-18^, *SMARCA4*^*11,13*^ and *TMEM106B*^*17*^, and antiviral genes such as *DAXX*^12^, *CABIN1, HIRA and XRCC3*^*11,13*^, *PIAS1* and *2*^6^. Several top pro/antiviral genes are involved in chromatin remodelling, DNA repair and histone modifications (i.e. epigenetic modifications). Epigenetic regulation occurs at multiple levels, including through DNA methylation, histone modification, RNA interference, nucleosome remodelling and modulation of 3D chromatin structure, and contains almost all molecular mechanisms affecting gene expression in a reversible, transmissible, and adaptive way without altering the sequence of genomic DNA.

Epigenetic modifications constitute antiviral restriction used by host cells as an innate immune defence against viral DNA and RNA^19^, they are also involved in the innate immune response^20^ to SARS-CoV-2 infection. For example, chromatin remodelling complexes SWI/SNF contribute to the activation of interferon-stimulated gene promoters and are master regulators of gene expression more broadly^21^. DNA methyltransferase enzymes (DNMTs) have a role in epigenetics and change in chromatin structures^22^. Epigenetic repression (i.e. rapid loading of histones bearing heterochromatic marks) also represses viral gene expression in a tug-of-war that is only revealed when viral countermeasures are experimentally removed^19^. PML-NBs are critical proteins involved in orchestrating the epigenetic repression of foreign DNA and RNA. Some PML-NB components increase in response to interferon signalling, and others, including the histone chaperone complex Daxx-ATRX, which loads histone variant H3.3, suppress viral replication. DNA viruses, including herpesviruses and HBV, package and/or encode viral proteins that can overcome this suppression^19^, highlighting PML-NBs as critical structures in the epigenetic suppression of viral activity.

The arms race between virus and host involves epigenetic mechanisms, including those innate to the host, those hijacked by the virus, and in the case of SARS-CoV-2, contributed to by viral protein histone-mimic ORF8^9^. What is not clear is how limitations imposed by host germline risk affect the expression changes that occur with infection induced chromatin remodelling. Specifically, the single gene impact from variants associated with the severity of SARS-CoV-2 infection has not been investigated for tissue-specific, infection-induced chromatin conformation changes temporally. In this study, we analysed gene targets of SARS-CoV-2 risk variants at different time points in lung epithelial cells: 0 hours (uninfected), 8 hours post-infection (8hpi), and 24 hours post-infection (24hpi)^7^. We also analysed gene expression data from lung tissue^4^. Recent results^9,11^ suggest there may be an epigenetic competition between the host and SARS-CoV-2. Here we find that germline risk in the severe phenotype, coupled with the epigenetic impact of SARS-CoV-2, limits the cells‘ ability to carry out relevant antiviral activities. Critically, the gene targets we identified are associated with host chromatin modifications amplified by SARS-CoV-2-induced epigenetic remodelling. Finally, we identify how this SARS-CoV-2-mediated process in genetically susceptible individuals mimics processes in normal and pathologically aged cells^23^, which may partly explain the severe phenotypes observed in younger patients and the exacerbated symptoms in some older patients. The profile of infection-induced (i.e. driven by differential interactions; lost/gained across time) gene activity, we identify, couples host germline risk with SARS-CoV-2-induced host epigenetic modifications^9^ for the first time.

## Results

Ho *et al*.^7^ identified structural changes to chromatin organisation in response to SARS-CoV-2 infection in human lung epithelial cells expressing the ACE2 receptor (i.e. A549^ACE2^ cells). We hypothesised that these structural changes to chromatin organisation were associated with altered variant-gene mapping and temporally defined alterations in transcriptional activity within the infected cells. CoDeS3D^24^ integrates GWAS data with Hi-C and eQTLs to identify putative tissue-specific functional outcomes associated with physical connections between regulatory sites and genes. We used CoDeS3D to integrate information on SARS-CoV-2 risk (i.e. variants associated with severe and hospitalised phenotypes), chromatin conformation data captured from uninfected (0h) and infected A549^ACE2^ cells^7^ at 8hpi (early) and 24hpi (late) time points post-infection, and eQTL data from lung tissue^4^ (Figure 1a). The spatial-eQTLs captured targeting genes within lung tissue are herein called ‘SCeQTLs‘. Across the three time points (i.e. uninfected, 8hpi and 24hpi), we detected a total of 30,438 regulatory interactions from 7,419 SCeQTLs involving SNPs associated with the severe phenotype and 33,284 interactions from 7,788 SCeQTLs for the hospitalised phenotype (Figure 1b; Supplementary table 1). These regulatory interactions involved 362 and 445 genes for the severe and hospitalised phenotypes, respectively (Figure 1b; Supplementary table 1). There was significant SCeQTL overlap between the hospitalised (76.5%) and severe (80.3%) phenotypes (Supplementary table 1), consistent with the observed overlap in SNPs associated with the SARS-CoV-2 severe and hospitalised severity phenotypes (Supplementary figure 1a). CoDeS3D identified 59 of the 95 genes within the COVID-19 HGI^1^ severe gene set (Supplementary table 2a).

**Figure 1.**
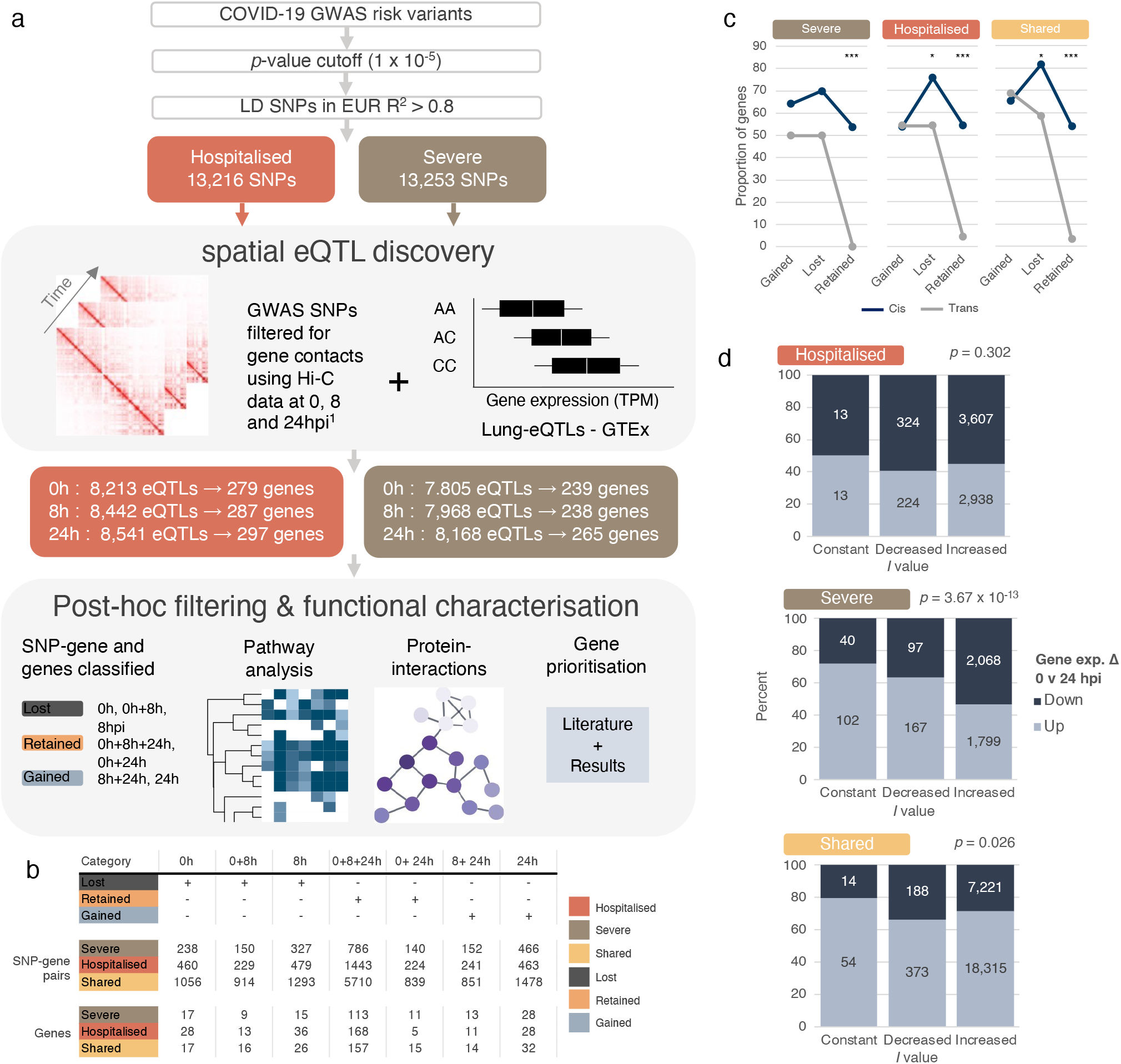
SARS-CoV-2 reprogrammes gene expression, including altering targets of SCeQTLs associated with infection severity. **(a)** Computational pipeline. The Codes3D pipeline generates a set of target genes associated with the severe and hospitalised phenotypes. SNPs obtained from COVID-19 HGI^1^ (Supplementary table 1) were screened through A549^ACE2^ cell Hi-C data from Ho *et al*. (2021)^7^ generated before and after SARS-CoV-2 infection (i.e. 0h, 8h and 24hpi) to identify *cis* (< 1 Mb), *trans* (> 1 Mb) and trans-interchromosomal SNP-gene interactions. eQTL effects were identified by testing the SNP-gene pairs against lung tissue in the GTEx database (version 8)^4^. SNP-gene pairs were classified according to time and retention following SARS-CoV-2 infection. The resulting statistically significant (FDR ≤ 0.05) SARS-CoV-2-specific genes (spatial lung eQTL-gene pairs, herein: SCeQTLs) from both the hospitalised and severe phenotypes were assessed for protein-protein interactions using the STRING^28^ database. All genes were assessed using g:Profiler^26^ pathway analysis (KEGG, REAC, HP and WP) tool to obtain gene ontology terms. A literature search further prioritised genes and the results found here (Supplementary table 4). **(b)** Count of SNP-gene pairs and SCeQTL-gene targets by time/category. **(c)** Proportion of genes that are lost/gained/retained from *cis* or *trans* interactions across shared/severe/hospitalised phenotypes. A two-sided test of proportions for equality was applied between *cis* and *trans*. **(d)** *I* value was calculated per gene based on the change in number of interactions over time. The proportion of interactions based on its effect (i.e. up/down regulation) by *I* value is shown. *p*-values were obtained from Fisher‘s test on 3×2 contingency tables (Supplementary table 3). *p<.05, **p<.01, ***p<.001.

Chromatin domains and loops are deregulated in SARS-CoV-2 infected A549^ACE2^ cells^7,8^. We hypothesised that these chromatin alterations would result in dynamic infection-associated changes in SCeQTLs and target genes. We classified SCeQTL-gene interactions unique to the severe and hospitalised phenotypes and those that were shared between the phenotypes as being “gained”, “lost”, or “retained” according to their dynamics upon infection. 484, 677 and 2,488 SeQTL-gene interactions were lost across the time course within the severe, hospitalised, and shared groups, respectively. 672, 963 and 5,326 SeQTL-gene interactions were retained, and 345, 427 and 1,825 SeQTL-gene interactions were gained (Figure 1b; See Methods). Collectively, these differential interactions account for 55%, 54% and 45% of the SeQTL-gene interactions within the severe, hospitalised, and shared groups, respectively (Supplementary figure 1e). One hundred sixty-five shared, 34 severe and 82 hospitalised genes were retained following infection (Supplementary figure 1d).

The proportions of *cis* and *trans*-regulated genes that are lost (hospitalised p = 0.0232; shared *p* = 3.72 × 10^−3^) and retained (shared p = 2.28 × 10^−7^; hospitalised p = 2.51 × 10^−6^; severe *p* = 9.87 × 10^−5^) significantly differed for the severe and hospitalised associated SCeQTLs (Figure 1c). *Trans* interactions were particularly dynamic. For example, of the forty-eight genes targeted by *trans*-only SCeQTLs (Supplementary figure 1c), forty were lost or gained with increasing time of infection. Notably, there was a significant increase in *trans-*acting SCeQTLs over the infection time course (Supplementary figure 1b).

To examine gene-level changes, we calculated the difference in the number of interactions for each gene targeted by a SCeQTL in uninfected cells and at 24hpi. The interactions were categorised based on constant, increased, or decreased numbers over the infection time course. There is a significant association between the difference in interaction numbers and gene transcript levels (i.e. up or down-regulation) for genes with SCeQTLs that were unique to the severe (Fisher‘s test *p* = 3.67 × 10^−13^) or shared between the severe and hospitalised (Fisher‘s test *p* = 0.026) phenotypes (Figure 1d). Across all interaction categories, the most significant effects in the shared group were associated with the upregulation of the target gene (Figure 1d; Supplementary table 3). For hospitalised genes with constant interactions (i.e. SCeQTLs equalled the same number over time), there was an equal association with up and down-regulation of the target gene transcript, in contrast to severe and shared phenotypes where constant interactions were proportionally associated with up-regulation (Figure 1d; Supplementary table 3).

Similarly, hospitalised genes with decreases in the number of SCeQTLs over time tended to be associated with downregulation of the gene transcript levels, while severe and shared genes in the same interaction category were associated with upregulation (Figure 1d; Supplementary table 3). Finally, hospitalised genes with SCeQTLs that increased over the infection time course were proportionally most associated with downregulation of the gene transcript levels, in contrast to upregulation observed in shared only-genes with the same interaction category (i.e. increased) (Figure 1d; Supplementary table 3). Overall, this suggests a different pattern of regulatory control of genes in the hospitalised phenotype.

### The regulation of established risk genes is differentially impacted by variants associated with hospitalisation or severe phenotypes across the SARS-CoV-2 infection time course

We hypothesised that the effect of chromatin remodelling following SARS-CoV-2 infection would implicate phenotype-specific transcriptional changes to established risk genes. Gene prioritisation was undertaken using two approaches, (1) SARS-CoV-2 associated host genes identified by functional studies (i.e. CRISPR screens)^11,13-18^ and (2) surfactant metabolism genes, which are expressed at lower levels in COVID-19 patients with ARDS^25^ (Figure 2; Supplementary table 4a). Genes within the HLA locus were excluded from this analysis.

**Figure 2.**
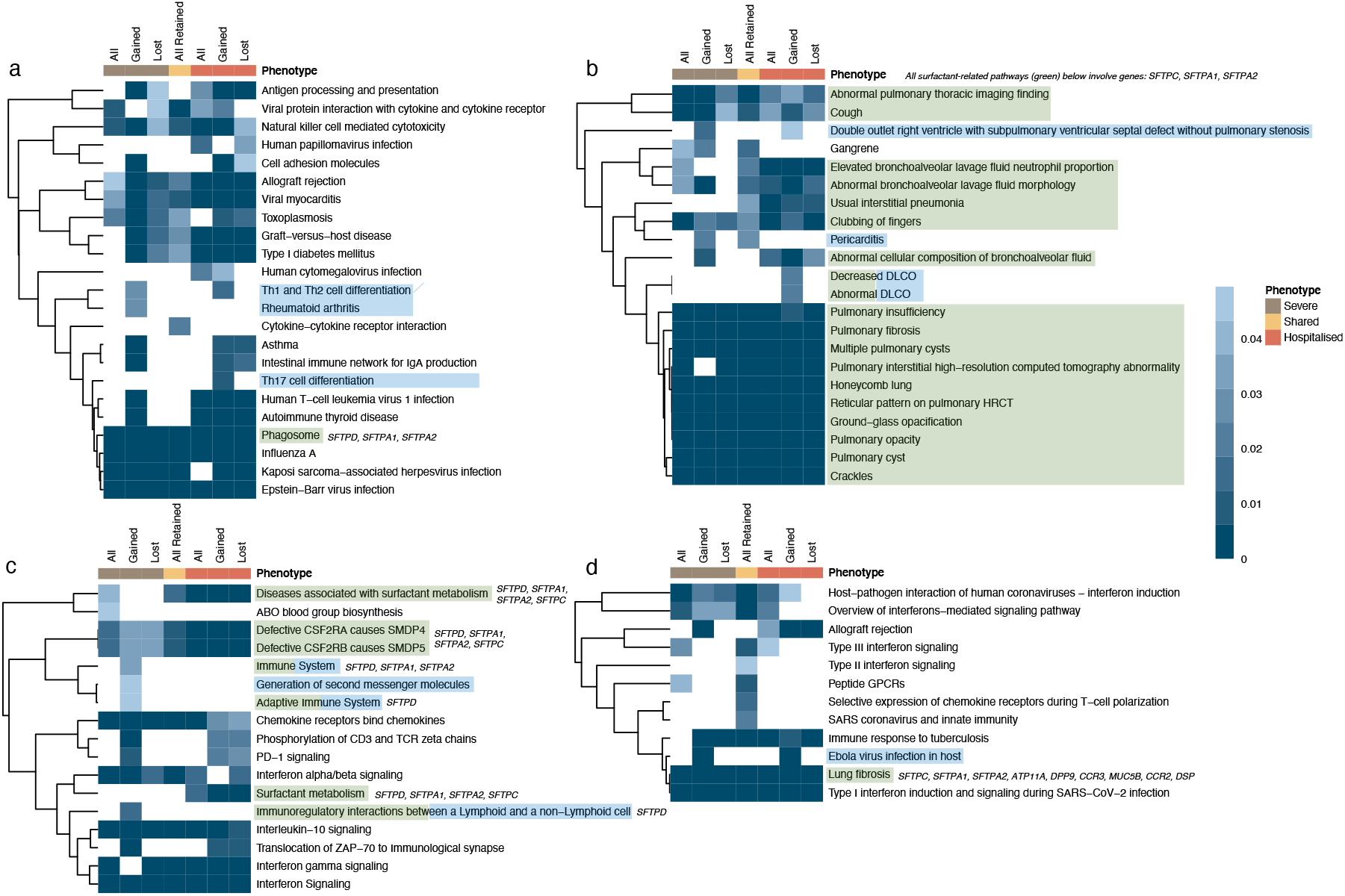
Gained and surfactant gene sets alter pathways associated with infection severity. Pathway analysis of all genes with transcript levels associated with SARS-CoV-2 risk variants over time. Pathways were categorised by gained, lost, and retained gene sets across both phenotypes. All retained = retained genes shared between both phenotypes, all = phenotype-specific retained genes and all retained, lost = all, and phenotype-specific lost genes gained = all, and phenotype-specific gained genes. **(a)** KEGG **(b)** Human Phenotype Ontology **(c)** Reactome and **(d)** Wiki Pathways databases were queried via g:Profiler^26^. K-means clustering was conducted on *p*-values. The most significant tend to be clustered around pathways driven by retained genes. Enrichment in pathways driven by surfactant genes is highlighted in green, and the surfactant genes involved are listed next to each pathway (Supplementary table 2a). We have highlighted all genes involved in lung fibrosis (*p* range = 6.98 × 10^−4^ to 2.89 × 10^−3^) due to its role in SARS-CoV-2 mortality^37^. We identified enrichment in processes that included antiviral programming (i.e. interferon response and signalling, chemokine signalling and lung function/pathologies). The gained category reflects pathways where the inclusion of eQTL gene targets that are expressed only in 24hpi and 8 + 24hpi drive the pathway enrichments. Among the 11 gained pathways (highlighted in blue), pericarditis (*p* = 0.024 and 0.029; severe and all retained, respectively), DLCO decrease (*p* = 0.019) and abnormal (*p* = 0.019) and Th17 (*p* = 0.007), Th1 and Th2 (*p* = 0.030 and 0.012; severe and hospitalised, respectively) cell differentiation support epidemiological evidence^56-58^ for clinically relevant phenotypes associated with SARS-CoV-2 (Supplementary table 2b).

Pathway analysis of all genes with transcript levels associated with SARS-CoV-2 risk variants across time identified enrichment in 28 processes that included surfactant metabolism genes (Figure 2; Supplementary table 4a). G:Profiler^26^ results were refined to include only the pathway databases results (i.e. WP, HP, REAC and KEGG), gene ontology terms were removed. Several pathways driven by surfactant genes are involved in functions of the immune system, i.e. phagosome (*p* range = 3.90 × 10^−5^ to 3.51 × 10^−3^), immune system (*p* = 0.03), adaptive immune system (*p* = 0.04), immunoregulatory interactions between a Lymphoid and a non-Lymphoid cell (*p* = 0.01). The remaining are directly involved in pathways covering functions/diseases of the lung. For instance, lung fibrosis (*p* range = 6.98 × 10^−4^ to 2.89 × 10^−3^), cough (*p* range = 0.001 to 0.037), and diseases associated with surfactant metabolism (*p* range = 9.77 × 10^−5^ to 0.04) (Supplementary table 4a). Whilst many of the pathways involve the same few surfactant genes (i.e. *SFTPD, SFTPC, SFTPA1, SFTPA2*; Figure 2), indicating the same pathways with different names, other genes are driving these enrichments (Supplementary table 4a).

The surfactant genes (*i*.*e. SFTA2, SFTPA1, SFTPA2, SFTPC, SFTPD and SFTPD-AS1*) encode components of the lung‘s innate immune system with roles in the maintenance of healthy lung function and clearance of pathogens^27^. The surfactant genes are primarily enriched in gained and retained pathways (Figure 2). Consistent with our hypothesis, the regulatory profiles of the surfactant genes (*SFTA2, SFTPA1, SFTPA2, SFTPC, SFTPD and SFTPD-AS1*) are specific to both the time post-infection and the genotypes associated with severe or hospitalisation (Figure 3). Most SCeQTLs associated with hospitalisation targeting *SFTPC* in the uninfected cells were retained across the infection time-course, like the hospitalised and severe *SFTPD*-SCeQTLs (Figure 3). SCeQTLs of the remaining surfactant genes (i.e. *SFTA2, SFTPA1, SFTPA2* and *SFTPD*-*AS1*) were lost and gained across the infection time-course in patterns that were specific to the hospitalised and severe phenotypes. For example, the *SFTA2*-SCeQTLs mainly were shared between the hospitalised and severe phenotypes at 24hpi. Notably, 8 out of 9 of the SFTA2-SCeQTLs at 24hpi were correlated with a decrease in transcript levels, and these SCeQTLs were gained over time. By contrast, *SFTPA1*-SCeQTLs mainly were (4 of 5) lost over the infection time-course, with only one SeQTL that correlates with an increase in *SFTPA1* transcript levels being present in both the hospitalised and severe phenotypes at 24hpi.

**Figure 3.**
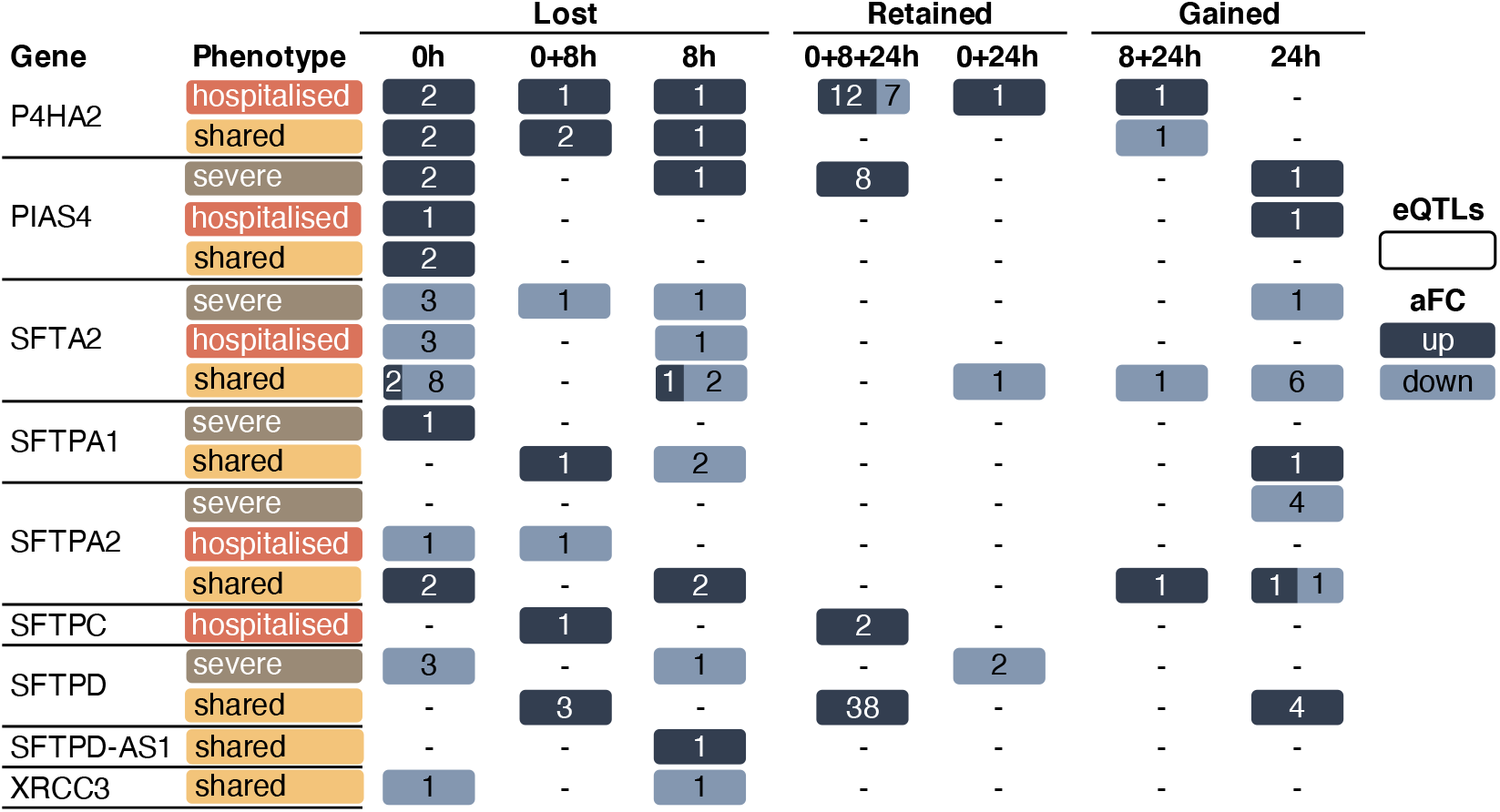
SCeQTL patterns associated with surfactants and other prioritised genes. SCeQTLs regulating the expression of prioritised genes are shown to be phenotypically distinct and governed by temporality. *P4HA4* is driving the association with pericarditis (Figure 2) and is preferentially upregulated in the hospitalised phenotype compared to severe (1 retained SCeQTL is downregulating). *PIAS4* shows more activity in the severe phenotype. Most regulatory activity to *SFTPA1* is lost. *SFTPA2* activity has a specific pattern of up/down-regulation associated with severe. Up regulatory activity associated with *SFTPC* is lost in the hospitalised phenotype. Across both phenotypes, *SFTPD* is predominantly being upregulated, with 2 retained SCeQTLs from severe downregulating. Across both phenotypes, *SFTPD-AS1* and *XRCC3* activity is lost (Supplementary table 1). eQTL; expression quantitative trait loci.

Amongst the seven *SFTPA2*-SCeQTLs at 24hpi, those associated with the severe phenotype predominantly correlated with decreases in transcript levels (i.e. 5:2 decrease: increase). By contrast, the hospitalised *SFTPA2*-SCeQTLs at 24hpi tended to increase transcript levels (2:1 ratio increase: decrease).

*SFTPC* and *SFTPD* were targeted by SCeQTLs that mainly were retained across the infection time course. For example, *SFTPC* transcript levels were upregulated by 2/3rds of the hospitalised-only SCeQTLs associated with this gene over time. Finally, 38 of the 45 *SFTPD*-SCeQTLs shared between hospitalised, and severe phenotypes were retained across the infection time course. Interestingly, the *shared SFTPD-SCeQTLs* correlated with increases in the transcript levels. By contrast, those *SFTPD*-SCeQTLs associated explicitly with the severe phenotype correlated with decreases in transcript levels. Notably, the only SCeQTL captured targeting *SFTPD-AS1* was associated with increased transcript levels and was lost at 8 hpi. *SFTPD* is the only ‘prioritised‘ surfactant gene that overlaps the COVID-19 HGI^1^ targets (Supplementary table 2a).

### Changes to the regulation of chromatin remodelling and transcription control genes are associated with infection severity

*XRCC3* (Chr 14) is an antiviral gene^11,13^ identified as a SARS-CoV-2 risk gene^11^. Two eQTLs were captured targeting *XRCC3* transcript levels in both the severe and hospitalised phenotypes. Notably, these SCeQTLs were correlated with reductions in *XRCC3* transcript levels, and both were lost by 24hpi, and the gene was no longer expressed (Figure 3; Supplementary table 1).

We hypothesised that the proteins encoded by the SCeQTL targeted genes would work together in shared biological pathways. We queried the STRING^28^ protein-protein interaction database to identify functionally related gene protein product clusters. The largest cluster of interacting genes (n = 40) included genes that are involved in chromatin remodelling (i.e. *SMARCA4, NCOR1, DNMT3A, DAXX, DNMT1, SETDB1*, and *CHD7*; Figure 4a; Supplementary figure 2). *SMARCA4* is widely recognised as being second to only *ACE2* as the most pro-SARS-CoV-2 viral gene^11,13^. SCeQTLs were captured targeting *SMARCA4* in both the hospitalised and severe phenotypes (Figure 4b; Supplementary table 1). However, the gain, loss, and retention patterns of these captured SCeQTLs differ in a phenotype-specific manner across the infection time course (Figure 4b; Supplementary table 1). The only SCeQTL captured upregulating *SMARCA4* was associated with the hospitalised phenotype in the uninfected cells – this SCeQTL was lost by 8hpi. Notably, there were 44 severe phenotype-associated SCeQTLs captured targeting *SMARCA4* at 24hpi. These 44 SCeQTLs were all correlated with reductions in *SMARCA4* transcript levels. By contrast, whilst it was also correlated with a decrease in *SMARCA4* transcript levels, there was only one hospitalised phenotype associated *SMARCA4*-SCeQTL at 24hpi (Figure 3b). We observed that the hospitalised/shared *SMARCA4*-SCeQTLs are located in more distal loci than the SCeQTLs associated with the severe phenotype (Figure 5a). These results are consistent with observations of proviral effects upon downregulation of SMARCA4^11^ and suggest the existence of a positive feedback loop involving SNPs associated with the severe phenotype in response to SARS-CoV-2 infection.

**Figure 4.**
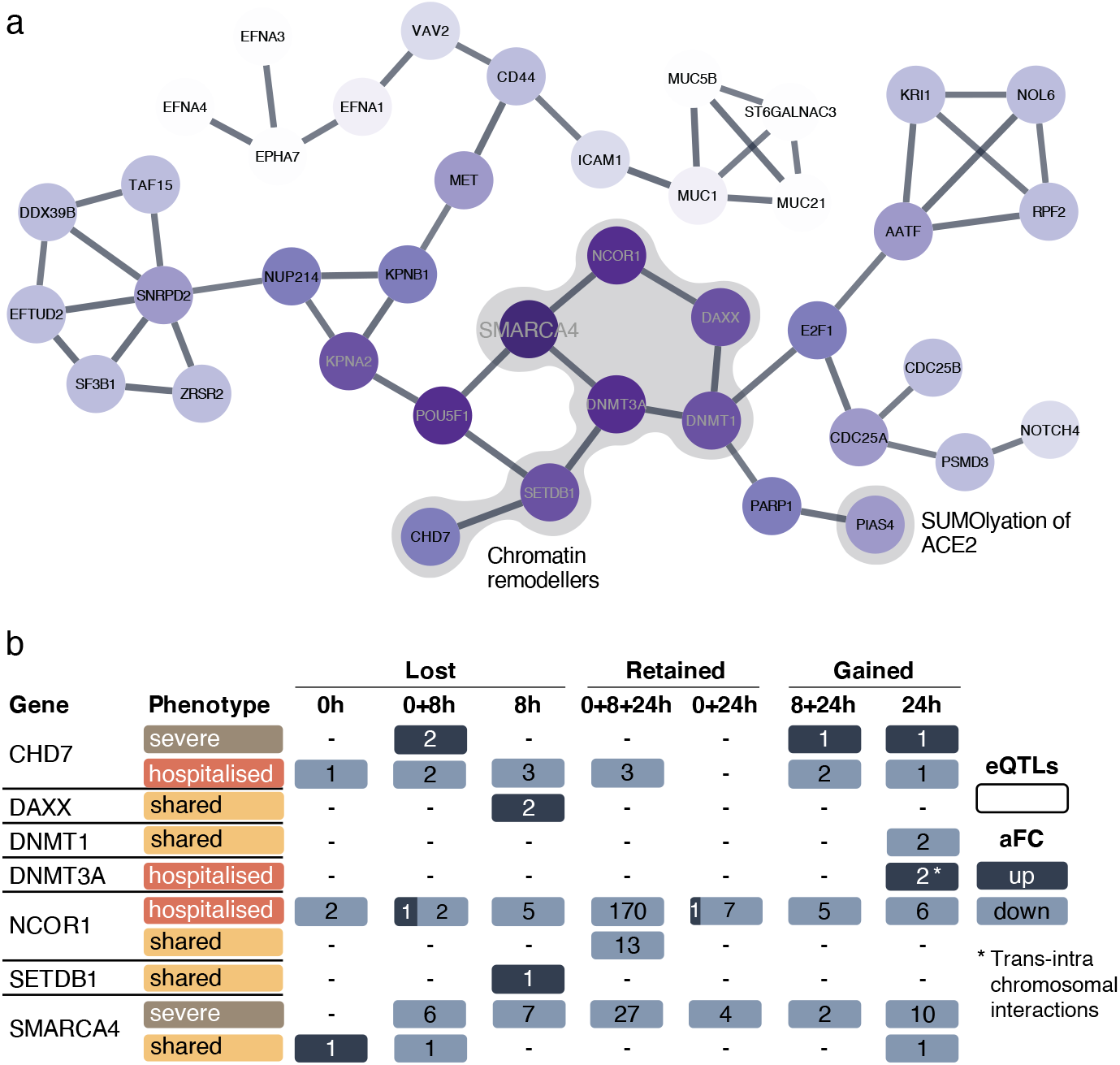
Chromatin remodellers form the largest protein cluster, with phenotypically divergent effect sizes. **(a)** The largest protein cluster from the entire STRING^28^ protein interaction network of genes (severe = 362, hospitalised = 445; Supplementary figure 2). SMARCA4 and its 6 close neighbours are all involved in chromatin remodelling. PIAS4 is also highlighted for its critical role in SUMOlyation of the ACE2 receptor^39^. **(b)** SCeQTLs regulating the expression of genes from (a) show unique phenotypic patterns with infection-induced alterations to chromatin, changing the number of interactions. aFC; allelic fold chang;

**Figure 5.**
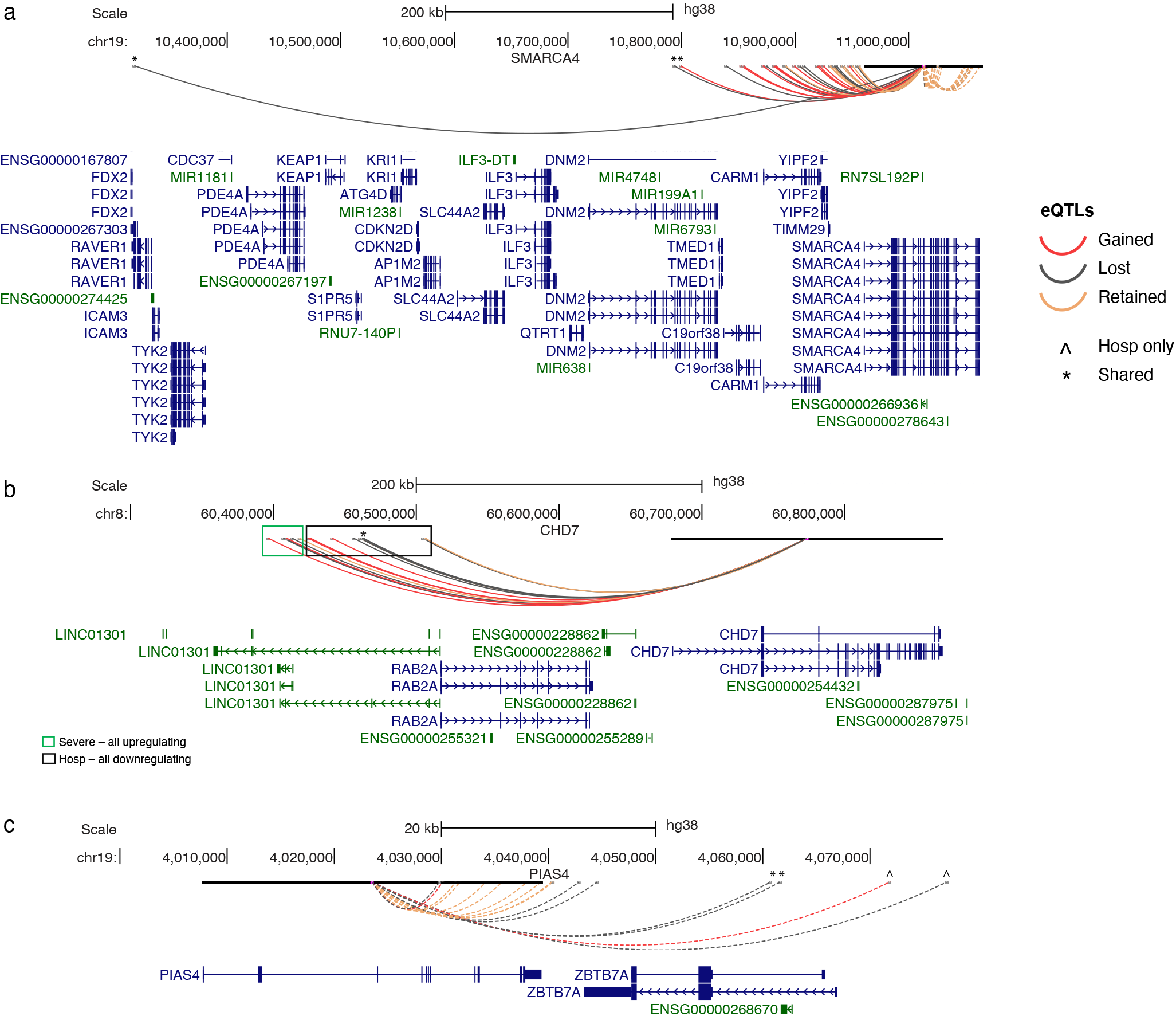
Genes that are phenotypically shared have distinct regulatory patterns. Genome tracks show SCeQTLs regulating *PIAS4* and chromatin remodellers, *SMARCA4* and *CHD7*.Hospitalised and shared eQTLs are annotated. Unannotated eQTLs are associated with the severe phenotype. There are defined blocks of SCeQTLs by phenotype. **(a)** *SMARCA4* highlights more distant SCeQTLs are shared, with only 1 retained in the hospitalised phenotype at 24h **(b)** *CHD7* SCeQTLs associated with severe in the more distant block all downregulate, whereas hospitalised SCeQTLs all upregulate. **(c)** Only 1 SCeQTL regulating *PIAS4* associated with the hospitalised phenotype is retained at 24hpi; the remaining are associated with severe. A similar pattern is observed in (a).

Among the other chromatin remodelling enzymes, *CHD7* is notable as the SCeQTLs that are gained, lost, and retained across time have opposite effects, up-regulatory and down-regulatory transcript levels in the severe and hospitalised phenotypes, respectively (Figure 4b). Notably, two severe phenotype-associated *CHD7*-SCeQTLs are lost, and two are gained across the infection time course. By contrast, 3 of 5 hospitalised associated *CHD7*-SCeQTLs are retained from the uninfected cells to 24hpi. As observed for *SMARCA4*, the *CHD7*-SCeQTLs were arranged in two distinct blocks (Figure 5b). These results are consistent with SARS-CoV-2 infection-dependent changes in *CHD7* regulation interacting with inherited genetic variation to impact infection outcome.

DNMT1 and DNMT3A encode cytosine-5-methyltransferases responsible for maintaining DNA methylation and *de novo* DNA methylation of CpG structures in the human genome, respectively^29^. Neither *DNMT1* nor *DNMT3A* has SCeQTLs in uninfected cells. However, SCeQTLs targeting both genes are captured at 24hpi. Notably, the SCeQTLs for *DNMT1* downregulates transcript levels, while those for *DNMT3A* upregulate transcript levels (Figure 4b; Supplementary table 1). The *DNMT3A*-SCeQTLs were located > 1Mb from the gene body and are likely to be involved in fine-tuning the regulation of *DNMT3A*. The upregulation of DNMT3A expression over the infection time course appears to be consistent with observations that amongst SARS-CoV-2 infected patients, worse outcomes were associated with a hyper-methylated status^30^.

The nuclear receptor corepressor 1 gene (*NCOR1*) ratio to the silencing mediator of retinoic acid and thyroid hormone receptor (*SMRT*) contributes to the fine-tuning of inflammatory versus tolerogenic balance^31^. Therefore, it is notable that most of the 210 SCeQTLs targeting down-regulation of *NCOR1* in uninfected cells were retained over the infection time course with SNPs associated with hospitalisation. Whilst only 13 of 210 SCeQTLs targeting the downregulation of *NCOR1* were captured in the severe phenotype and were retained out to 24hpi (Figure 4b; Supplementary table 1).

Daxx is an epigenetic repressor of viruses^32^, and a potent inhibitor of SARS-CoV-2 and SARS-CoV replication in human cells^12^. Daxx forms a complex with ATRX and is mainly known for its antiviral activity against DNA viruses replicating in the nucleus^19^. However, DAXX can also restrict SARS-CoV-2 by rapid re-localisation to the cytoplasmic viral replication sites^32^. Therefore, it is notable that two SCeQTLs that were captured and associated with the upregulation of DAXX transcript levels in uninfected cells were lost by 8hpi. These eQTLs were shared between the hospitalised and severe phenotypes (Figure 4b; Supplementary table 1). This is consistent with SARS-CoV-2 mediated chromatin remodelling mediating down-regulation of Daxx expression during the infection time course.

The tissue specificity of both eQTLs and chromatin structure is widely recognised^4,33-35^. We parsed the SCeQTL-gene pairs associated with the chromatin remodelling cluster (Figure 4a) through the GTEx catalog^4^ to determine their tissue specificity (Figure 6; Supplementary table 5). Of 293 SCeQTL-gene pairs associated with regulating the seven chromatin remodelling genes (i.e. *SMARCA4, NCOR1, DNMT3A, DAXX, DNMT1, SETDB1*, and *CHD7*), 278 (94.8%) were present in ‘cells cultured fibroblasts‘. Superficially, this is consistent with observations that pathological fibroblasts^36^ contribute to rapidly ensuing pulmonary fibrosis in COVID-19^37^.

**Figure 6.**
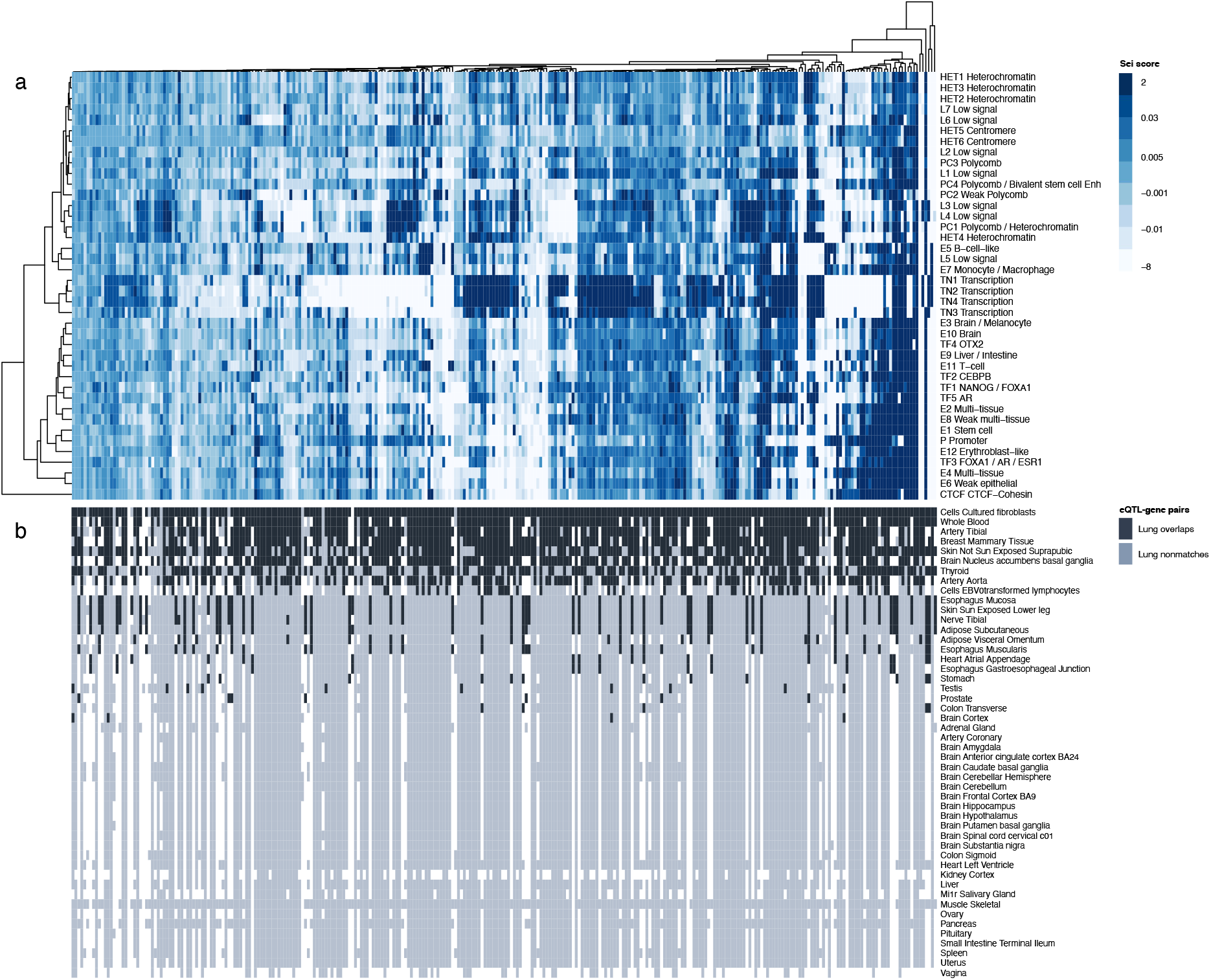
Lung-SCeQTLs regulating chromatin remodellers also target fibroblast cells, with predicted regulatory activities driving the overlaps. 293 lung-SCeQTLs associated with chromatin-modifying genes were screened for eQTL-gene pair matches in GTEx^4^ tissues (n = 48) to determine tissue-specificity. The same SCeQTLs were parsed through Sei^38^ to determine predicted regulatory functions. Positive Sei scores indicate an increased sequence class activity by the alternate alleles. Each column represents 1 lung-SCeQTL, and rows represent **(a)** predicted Sei regulatory functions for each SCeQTL and **(b)** whether the lung-SCeQTL-gene targets are the same in other GTEx tissues. Lung-SCeQTL-gene pair matches from CoDeS3D are coloured navy, and nonmatches are coloured light blue. The heatmap is ordered by the number of SCeQTL-gene pair overlaps with the lung. There are 0 eQTL-gene pair overlaps in 26 tissues, 10 tissues < 50, and the remaining 12 tissues 50 – 293 (Supplementary table 5b). ‘Cells cultured fibroblasts‘ (n = 278), ‘whole blood‘ (n = 234) and ‘artery tibial‘ (n = 220) were the top 3 overlapping tissues. The greater the Sei score, the higher the confidence in predicted regulatory activity. There are distinct blocks of transcription and enhancer activity.

The effects of these SCeQTLs in ‘whole blood‘ (n = 234; 79.8%) and ‘artery tibial‘ (n = 220; 75.0%) were also largely conserved (Figure 6; Supplementary table 5b). Among the tissues where these effects were not conserved (i.e. eQTLs were identified but were not associated with the same lung gene transcripts), ‘muscle skeletal‘ (n = 280; 95.6%), ‘pancreas‘ (n = 250; 85.3%) and ‘adipose subcutaneous‘ (n = 244; 83.3%) (Figure 6b; Supplementary table 5b) were three tissues with the most eQTL activity. Sei^38^ predicted the regulatory activity of chromatin-SCeQTLs by integrating 21,907 chromatin profiles (transcription factor, histone marks, and chromatin accessibility profiles across a wide range of cell types). This identified clusters of enhancer activity and transcription driven mainly by the transcription 1-4 profiles and aligned with clusters of lung-SCeQTLs-gene targets from other tissues in GTEx (Figure 6a). Notably, the transcription 1-4 profiles identified by Sei are primarily defined by H3K36me3 and the patterns of RELA, TAL2, SOX10, HNF4A, TP53, AR, CEBPB, OTX2 and NANOG binding in different cell types, and an absence of CTCF binding in the A549 lung epithelium. ^38^

### Germline risk in PIAS4 involved in SUMOylation of ACE2

The E3 SUMO ligase PIAS4 promotes the SUMOylation and stabilisation of ACE2. PIAS4 knockouts have reduced SARS-CoV-2 infection^39^. Germline risk variants associated with both the hospitalised and severe phenotypes were captured as SCeQTLs targeting *PIAS4* transcript levels (Figure 3). However, the SCeQTLs associated with the severe phenotype were more dynamic over the infection time course when compared to the hospitalisation phenotype. Notably, from the uninfected cells to 24hpi, 8 *PIAS4*-SCeQTLs from the severe phenotype were upregulating *PIAS4*. By contrast, only 1 SCeQTL from the hospitalised phenotype was captured targeting upregulation of the *PIAS4* gene transcript at 24hpi. As observed earlier, the hospitalised and shared SCeQTLs were located within blocks more distal to the *PIAS4* gene (Figure 5c). Notably, 3 out of 4 of these distally located hospitalised SCeQTLs were lost over the infection-time course. The observation that all of the SCeQTLs captured targeting *PIAS4* were associated with increases in transcript levels is consistent with observations by Jin *et al*.^39^ that suggest a more significant burden of *PIAS4* in the severe phenotype, with greater activity retained at 24hpi.

## Discussion

We combined temporal data from SARS-CoV-2 stimulated cells^7^, and a spatial-eQTL analysis in lung tissue to identify gene regulatory changes that occur across the SARS-CoV-2 infection time course. Our results provide evidence for the direct effects of infection-induced chromatin remodelling on regulatory impacts of inherited germline variants that are associated with severity of SARS-CoV-2 infection. The results for the effect of eQTL target genes, such as *PIAS4, SMARCA4, XRCC3* and *DAXX*, on the severity of infection are supported by published functional assays^9,11,13-18^. The severe phenotype-associated eQTLs that were captured targeting genes involved in chromatin remodelling (i.e. *SMARCA4, NCOR1, DNMT3A, DAXX, DNMT1, SETDB1, CHD7)*, surfactant metabolism (i.e. *SFTA2, SFTPA1, SFTPA2, SFTPC, SFTPD and SFTPD-AS1)*, and viral responses (e.g. *XRCC3*^11,13^ and *PIAS4*^39^) form and break in an infection time-course dependent manner that mimics positive feedback loops connecting germline variation with the process of viral infection and replication.

The analysis of SARS-CoV-2-stimulated cells we performed uncovered previously undescribed lung-specific gene targets associated with severe COVID-19. However, this study has several limitations. 1) We were limited to analysing common genetic variants (MAF ≥ 0.05). Including rare variants with larger effect sizes may impact additional genes with more significant phenotypic consequences. We have, however, identified a link in gene deletion studies and their control elements (i.e. common SARS-CoV-2 risk variants highlighted here), which converges two methods for functional annotation, reinforcing the potential significance of these genetic variants at a population level. 2) The protein interaction networks depended on known and predicted protein interaction data (including direct and indirect functional associations) within the STRING database^28^. Likely, this dataset does not capture all biologically relevant protein interactions. Finally, 3) we did not capture the spatial genome organisation [e.g. Hi-C data] and gene expression data from identical samples. Therefore, inter-sample variation between the different datasets will impact the analysis. Notwithstanding these limitations, we have systematically identified the lung-specific regulatory role of variants associated with hospitalisation and severe SARS-CoV-2 infection and captured how these interactions change over time under infection conditions.

COVID-19 occurs within physiological limits that result from the impact of the patient‘s germline and somatic variants impacting environmental signals over a lifetime. As such, the germline variants significantly contribute to the limits of how the patient‘s cells and tissues respond to infections, including SARS-CoV-2. These contributions need not be fixed by the developmental state of the cell, as evidenced by the interactions between the germline variants and SARS-CoV-2 infection-induced chromatin remodelling on gene regulation that we identified. Of course, the remodelling occurs in all genotypes and is not limited to those prone to more severe infection. However, the presence of germline variants associated with severity of infection within control elements highlights a) those regulatory interactions that change and b) dynamic changes (i.e. increases or decreases in expression) that contribute to infection severity. For example, pulmonary surfactants are a complex of lipids and proteins that enhance pathogen clearance and regulate adaptive and innate immune-cell functions^27^. SARS-CoV-2 targets alveolar type-II cells, the lung cells that produce surfactant^40^. At the same time, surfactant levels are markedly reduced in the subset of COVID-19 patients who present with ARDS^25,41^. Therefore, the observation that germline risk variants impact the temporal pattern of surfactant gene regulation across the SARS-CoV-2 infection time-course identifies critical targets (e.g. *SFTPD, SFTPA2, SFTA2*) for therapeutic intervention through the restoration of ‘typical‘ expression levels. In support, exogenous surfactant treatment in some patients with COVID-19 experiencing respiratory failure was found to be the catalyst for the successful extubation and clinical improvement of the patient^42^. Finally, several clinical trials are currently examining the use of exogenous surfactants to treat SARS-CoV-2-induced ARDS, highlighted in this review (Herman *et al*.^43^).

Chromatin remodelling impacts gene regulation and expression by providing the transcription machinery with dynamic access to genes and control elements (via nucleosome movement) to an otherwise tightly packaged genome. There is significant chromatin remodelling across the SARS-CoV-2 infection time course^7,8^. We provide evidence that this remodelling is enhanced in genotypes susceptible to severe SARS-CoV-2. This results from regulatory changes to chromatin and DNA-associated gene targets we have identified and acts in a positive feedback loop associated with severity. Among the chromatin-associated target genes we identified, the proteins of *CHD7, DAXX*, and *SMARCA4* can all function as remodellers via editing (incorporating histone H3.3)^21^. We identified a phenotypic switch associated with the regulation of *CHD7. SMARCA4* has more proviral regulatory activity in the severe phenotype, and the up-regulation of *DAXX* is lost due to SARS-CoV-2-induced chromatin changes - all contributing to a more proviral environment. The importance of these host restrictions in SARS-CoV-2 is supported by recent results from a meta-analysis of SARS-CoV-2 CRISPR experiments showing H3.3 chaperone complexes, *HIRA* and *CABIN1* highly significant for antiviral activity across all 6 cells lines included^13^. *SMARCA4* was also directly identified as being proviral^11^. In addition, viruses have adapted ways to overcome these host restrictions. For example, HIV utilises host ATP-dependent chromatin remodelling factors to displace nucleosomes, thereby increasing DNA accessibility for HIV integration^44^. Herpesviruses and HBV produce viral proteins targeting the PML-NB components such as Daxx-ARTX^19^. SARS-CoV-2 mimics a histone protein that effectively suppresses type 1 interferon-responsive antiviral genes^9^. While well-established interferon-associated gene targets (i.e. *IL10RB, IFNAR2;* Supplementary table 1) associated with severe COVID-19^1-3^ likely contribute to limiting the cell‘s ability to respond effectively to SARS-CoV-2 under the conditions discussed. Evidence supports a mechanistic role for these genes in the pathogenesis of severe COVID-19.

CRISPR assays^11,13-18^ support the putative roles for *XRCC3* (antiviral) and *SMARCA4* (proviral)^11,13^, *PIAS4* (antiviral)^39^ and *DAXX* (antiviral)^12^ that we identified as lung-specific regulatory targets and changing across the SARS-CoV-2 infection time course. Critically, we show that in most tissues we compared with (discussed below), there is no overlap in eQTL targets associated with these genes; therefore, contextually, these effects are lung-specific and will be different in other cells/tissues. Among the gene targets, *XRCC3, SMARCA4* and *DAXX* are directly involved in transcriptional control and chromatin remodelling. At the same time, the PIAS4 protein is the crucial SUMO ligase prompting the SUMOlyation and stabilisation of ACE2^39^, which is essential for SARS-CoV-2 replication. In line with the results of Jin *et al*.^39^ which showed a more severe infection with higher levels of PIAS4, we found the severe phenotype to have more up regulatory activity compared with one only up regulatory eQTL gained in the hospitalised phenotype across the infection time course. This finding is limited to the late stage of infection, i.e. 24hpi, whereas severe is not. Therefore, the physiological limits imposed by the severe-only eQTLs targeting *PIAS4* are not limited to a particular phase of infection. We have previously identified *SMARCA4* as an eQTL target gene associated with the severe and hospitalised SARS-CoV-2 phenotypes^6^. However, the temporal data integration highlights the role of severe-only *SMARCA4* eQTLs in producing a more significant regulatory burden compared with the more distant and temporally-regulated SMARCA4 eQTLs in the hospitalised phenotype. The only eQTL upregulating *SMARCA4* is lost by 8hpi, which based on the evidence presented here, is associated with a loss of antiviral activity. *XRCC3*^11,13^ and *DAXX*^12^, both shared genes, provide an example of how the cell tries to reduce infection burden by expressing these antiviral genes; however, lose their interactions due to SARS-CoV-2 and, therefore, antiviral activity. *DAXX*, in particular, forms part of a well-established protein complex Daxx-ATRX, mainly known for its role in antiviral activity against DNA viruses^19^, but is shown to rapidly re-localise to the site of SARS-CoV-2 viral replication within the cytoplasm and represses transcription^32^. Loss of this activity was previously shown^32^ to reduce the cell‘s ability to block the effects of SARS-CoV-2. Whilst there may be regulatory activity to these genes not associated with infection severity, in people with this germline risk profile, the proviral environment is amplified directly by SARS-CoV-2. Finally, among shared genes with few proviral hospitalised-only eQTLs, such as *SMARCA4* and *PIAS4*, this highlights a scenario where limitations of the cell (i.e. germline risk) are less than the severe phenotype, whereby the direct effects of SARS-CoV-2 are reduced.

We observed a tendency for the distance between eQTLs, and their target genes to be associated with the hospitalised and severe phenotypes (e.g. *SMARCA4* and *PIAS4*). Despite variants in the hospitalised and severe phenotypes sharing target genes within lung tissue, we suggest that localised promoter elements are associated with severe cases because they are hitting the central control elements of the gene. In contrast, distal eQTLs associated with the hospitalised phenotype are involved in fine-tuning gene expression. It is trivial that the location of variants within conserved ‘core‘ promoter elements will impact gene regulation, whereas more distal eQTLs are likely to fine-tune regulation and have smaller effects^45^, these observations need to be confirmed for the situations identified in this manuscript.

Epigenetic modifications are part of a dynamic process involved in immune defence and antiviral restriction^19^. We identified *NCOR1* and DNMTs 1 and 3a as genes whose regulation dynamically changes with infection and is altered in people susceptible to hospitalisation and severe infection. DNMTs may influence antiviral responses^22^, via methylation-induced transcriptional suppression. By inhibiting DNMTs with decitabine (known to reduce levels of DNMT1 and DNMT3a), Hennessey *et al*.^22^ found that transcription of Toll-like receptor 3, a pattern recognition protein for the innate immune system, was significantly down-regulated. Administration of decitabine also accelerated the resolution of lung injury in a mice model of ARD via an increase in regulatory T cells^46^. The evidence suggests that the gained severe and hospitalised eQTLs that correlated with the downregulation of *DNMT1* may be protective, and the hospitalised-only eQTLs upregulating *DNMT3a* may contribute to a more serious clinical phenotype. This is an area of ongoing investigation, with one clinical trial studying the efficacy of decitabine in COVID-19-related ARDS^47^. *NCOR1* is a well-studied transcriptional corepressor complex that represses the function of nuclear receptors and thus regulates critical inflammatory and metabolic processes^48^. NCoR1 genetically inactivates HDAC3, which impairs the activation of pro-inflammatory genes in the hyperinflammatory response to LPS in macrophages^49^. Down-regulation of *NCOR1* and potential loss of the NCoR1 protein may remove the HDAC3-associated break on hyperinflammatory syndrome induced by SARS-CoV-2, associated with disease morbidity and mortality^50^. NCoR1 may form part of this aberrant process and potentially be targetable.

The transcriptional response to a defined stimulus can vary among cell types^34^. Here, we identified eQTLs correlated with the expression of its target genes in lung tissue. However, we also found that lung-eQTLs correlated to transcripts of the chromatin remodeller genes are more than 90% the same in fibroblast cells. These cells are involved in the repair and remodelling of the extracellular matrix. By producing ECM-remodelling enzymes and inflammatory cytokines, damage-responsive fibroblasts modify the lung microenvironment to promote robust immune cell infiltration at the expense of lung function^36^. Fibroblasts can also support viral replication and contribute to the inflammatory response in the lungs, while pathological fibroblasts^36^ contribute to rapidly ensuing pulmonary fibrosis in COVID-19^37^. The interaction between germline risk and SARS-CoV-2 in epithelial cells and lung tissue may also function similarly in fibroblasts. This presents an intriguing possibility that the interaction between germline risk and gene regulation we identified occurs in lung fibroblasts and contributes to COVID-19-induced pulmonary fibrosis. Confirming that these interactions occur in non-immortalised fibroblasts in response to SARS-CoV-2 infection-induced chromatin remodelling is essential for developing therapeutic interventions to reduce this complication.

Elderly populations are at higher risk of severe disease and death from SARS-CoV-2^51^. Epigenetic mechanisms, such as changes to chromatin architecture and global heterochromatin loss and redistribution, are characteristic features of aging^23,52^. Therefore, the infection-induced regulatory changes in chromatin and DNA-modifying gene expression we identified may lead to changes that mimic the epigenetic profile of aged cells. Several lines of evidence support this hypothesis, 1) In aged cells, the expression of DNMT1 is decreased, while DNMT3a is increased^52^. This matches the patterns observed from the risk variants targeting these genes in the severe and hospitalised phenotypes. 2) The remodelling complex SWI/SNF is required for co-expression of the telomere binding proteins, which are essential for maintaining telomere length and structure in human fibroblasts^52^, and inhibition of SMARCA4 prevented aging-dependant proteins shortening the lifespan in *Drosophila* Parkinson‘s models. This aligns with the SWI/SNF gene target (i.e. *SMARCA4* and *CHD7*) patterns observed in the severe phenotype. Our results may partly explain rare cases of young patients with severe disease while contributing to the burden in older adults.

Assigning target genes to GWAS-identified variants remains complex^53^, and is highlighted by the large difference (38%) in gene targets we mapped compared to the COVID-19 HGI consortium using the same risk variants. The tissue specificity of both eQTLs and chromatin structure is widely recognised^4,33-35^. The eQTL context-dependence (i.e. cell/tissue specificity and development state) has an effect. It is therefore not surprising we see this divergence, the COVID-19 HGI used methods that did not include tissue-specific gene expression, nor did they account for physical chromatin interactions between the risk variant and its predicted gene target. In addition, the overlaps we identified between the CRISPR screen results^11,13-18^ and CoDeS3D uncover the compound effect of host germline risk with SARS-CoV-2-specific pro/antiviral host physiological factors.

## Conclusion

In conclusion, we present evidence for the convergence of multiple gene regulatory mechanisms, including SARS-CoV-2-induced changes in chromatin organisation^7-9^, with predicted lung-specific transcription of pro/antiviral host factors associated with germline risk to severe infection. We show that several genes forming protein interactions are involved in critical aspects of the cell‘s antiviral response and are supported by functional assays of SARS-CoV-2 infection^11,13-18^. Two crucial findings are associated with our results: (1) several of the genes targeted by germline variants associated with the severity of SARS-CoV-2 infection are transcriptionally affected by host chromatin modification to epithelial cells under SARS-CoV-2 infection conditions. These include the chromatin remodeller, surfactant genes and genes involved in viral response, which may mimic positive feedback loops. (2) the interactions driving the regulation of these genes are phenotypically distinct, shown here to be more skewed to the severe phenotype regardless of sharing gene targets. Under normal conditions, these germline limitations may have minimal physiological impact. However, we show how under pathological conditions (SARS-CoV-2 infection), these limitations enhance the proviral environment in the lung via SARS-CoV-2-mediated chromatin modifications.

## Methods

### Identification of SNP-gene pairs following infection with SARS-CoV-2

Genome-wide association study (GWAS) data for SARS-CoV-2 clinical phenotypes was obtained from the Covid-19 Host Genetics initiative^2,3^ (COVID-19 HGI). Single nucleotide polymorphisms (SNPs) for the hospitalised versus population and severe (hospitalised AND death or respiratory support) versus population (*p*-value threshold of 1 × 10^−5^) cohorts were obtained from COVID-19 HGI release 7 (https://www.covid19hg.org/results/r7/; Supplementary Table 1). SNPs in linkage disequilibrium within 5,000 base pairs at an R^2^ threshold of > 0.8 from European populations were obtained from 1000 Genomes phase 3 data^54^. Full summary statistics and details from COVID-19 HGI are available at https://app.covid19hg.org/44.

### Assigning putative transcriptional functions to SARS-CoV-2 SNPs across time following infection

SARS-CoV-2-associated SNPs were analysed using CoDes3D^24^ to identify spatially constrained expression quantitative trait loci (eQTLs) and their target genes. Spatial connections for each SNP-gene pair were identified from Hi-C chromatin contact data derived from uninfected (0h) and infected (8h and 24hpi) A549^ACE2^ cells^7^ (GEO accession: GSE162612). To identify which SNPs, with minor allele frequency threshold of 0.05, are eQTLs, the SNP-gene pairs were used to query lung tissue within the GTEx database (version 8)^4^. Multiple testing was corrected using the Benjamini-Hochberg procedure^55^ (FDR < 0.05), and interactions were kept if the logarithm of allelic fold change (log_aFC) ≥ 0.05. eQTL and gene chromosome positions were annotated using the human reference genome GRCh38/hg19.

### Analysis of gene expression dynamics based on interactions over time

Based on a modified analysis within the Ho *et al*.^7^ paper, the SCeQTL-gene interaction data was curated based on assignment into three categories: lost (“0h” and “0h+8h”), retained (“0h+8h+24h” and “0h+24h”) and gained (“8h+24h” and “24h”), respectively, upon infection. We used the same interaction categories to assess the relationship between gene transcript levels and changes in the number of interactions. Additionally, we classified the SCeQTL-gene pairs into “down” (log2aFC < 0) and “up” (log2aFC > 0). For each gene, the change in the number of interactions *I* was computed as follows:

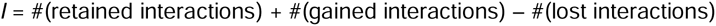

Genes were partitioned into three categories based on *I*: “reduced” (*I* < 0), “constant” (*I* = 0), “increased” (*I* > 0) (Supplementary table 3). We used Fisher‘s test on 3×2 contingency tables to assess significance. Results were presented as proportion plots with each combination of gene transcript and interaction category shown by phenotype; severe, hospitalised and shared.

### Pathway analysis

Gene Ontology (GO) and Kyoto Encyclopedia of Genes and Genomes (KEGG) pathway enrichment analysis was conducted using the g:GOSt g:Profiler (‘gprofiler2‘) R package. The Reactome (REAC), WikiPathways (WP) and Human Phenotype Ontology (HP) databases were included. Pathways and significant terms were selected based on the g:SCS default algorithm with the threshold of adjusted *p*-value < 0.05 (Supplementary table 4). Statistical significance was calculated considering only annotated genes of the hsapiens organism in the Ensembl database.

### Protein-protein interaction network analysis

Curated protein-protein interaction data were obtained from STRING (https://string-db.org). STRING was mined using lists of genes targeted by spatially constrained eQTLs and the following parameters: experiments, text mining, co-expression and databases, and species limited to “Homo sapiens”. The network was constructed with the highest evidence interaction score of ≥ 0.9. Singletons were removed from the network.

### Systematic prediction of sequence regulatory activities from SARS-CoV-2 eQTLs

Sei^38^ is a deep-learning-based framework for systematically predicting sequence regulatory activities and applying sequence information to understand human genetics data. SNPs from COVID-19 HGI severe and hospitalised phenotypes, along with all LD SNPs, were annotated with chromosomal position, reference (REF) and alternate (ALT) allele using the GTEx lookup table for all variants genotyped in GTEx, with chromosome positions, REF and ALT alleles, SNPs from dbSNP 151, GTEx variant IDs (constructed as chr_pos_ref_alt_build), and hg19 liftover variant ID, for all variants in release V8. Input eQTLs were queried for regulatory activities using the Sei web server (https://hb.flatironinstitute.org/sei/).

### Gene prioritization

Gene prioritisation was undertaken to determine the effect of regulatory changes over time. Data for gene prioritisation was obtained from two sources; (1) SARS-CoV-2 associated host genes identified by functional studies (i.e. CRISPR screens)^11,13-18^ and (2) surfactant metabolism genes (Figure 2; Supplementary table 4a). Genes within the HLA locus were excluded from this analysis. The COVID-19 HGI consortium GWAS target genes predicted from V2G, eGenes, coding variants, and distance (closest gene) were filtered by COVID-19 phenotype ‘critical illness‘ & ‘hospitalised‘. Ninety-five predicted gene targets from COVID-19 HGI^1^ hospitalised and severe datasets were extracted and screened for spatial lung-eQTL activity using CoDeS3D results (Supplementary table 2). Custom tracks (https://genome.ucsc.edu/cgi-bin/hgCustom) on the UCSC Genome Browser human genome assembly GRCh38/hg38 were used to visualise SCeQTLs. The GENCODE V41 track was also included. To identify lung SCeQTL-gene matches from the 7 clustered chromatin remodellers genes, GTEx (version 8) was queried for *cis* eQTLs-gene pairs in the remaining 48 tissues in the catalogue.

### Availability of data and materials

Codes3D - https://github.com/Genome3d/codes3d

GWAS data - https://www.covid19hg.org/results/r7/

Hi-C data - https://www.ncbi.nlm.nih.gov/geo/query/acc.cgi?acc=GSE162612

GTEx data - https://storage.googleapis.com/gtex_analysis_v8/reference/GTEx_Analysis_2017-06-05_v8_WholeGenomeSeq_838Indiv_Analysis_Freeze.lookup_table.txt.gz

## Data Availability

All data produced in the present work are contained in the manuscript, and all data and materials used in the manuscript can be accessed online here:
Codes3D - https://github.com/Genome3d/codes3d 
GWAS data - https://www.covid19hg.org/results/r7/ 
Hi-C data - https://www.ncbi.nlm.nih.gov/geo/query/acc.cgi?acc=GSE162612 
GTEx data - https://storage.googleapis.com/gtex_analysis_v8/reference/GTEx_Analysis_2017-06-05_v8_WholeGenomeSeq_838Indiv_Analysis_Freeze.lookup_table.txt.gz 

## Declarations

### Ethics approval and consent to participate

While deceased individuals do not require consent for research, GTEx consent is described in detail^4^. Ethics approvals for COVID-19 HGI GWAS data are also described in detail^2,3^.

### Consent for publication

Not applicable.

### Availability of data and materials

See methods.

## Author contributions

RKJ performed data analyses and interpretation, created figures, and wrote the manuscript. EG contributed to manuscript revision. JOS directed the study, contributed to data interpretation, and co-wrote the manuscript.

## Acknowledgements

The Genotype-Tissue Expression (GTEx) Project was supported by the Common Fund of the Office of the Director of the National Institutes of Health and by NCI, NHGRI, NHLBI, NIDA, NIMH, and NINDS. The authors thank the Genomics and Systems Biology Group (Liggins Institute, University of Auckland) for valuable discussions. The authors acknowledge the COVID-19 Host Genetics Initiative consortium for providing infrastructure and access to the SARS-CoV-2 GWAS meta-analysis data.

## Supplementary Figures

**Supplementary figure 1.**
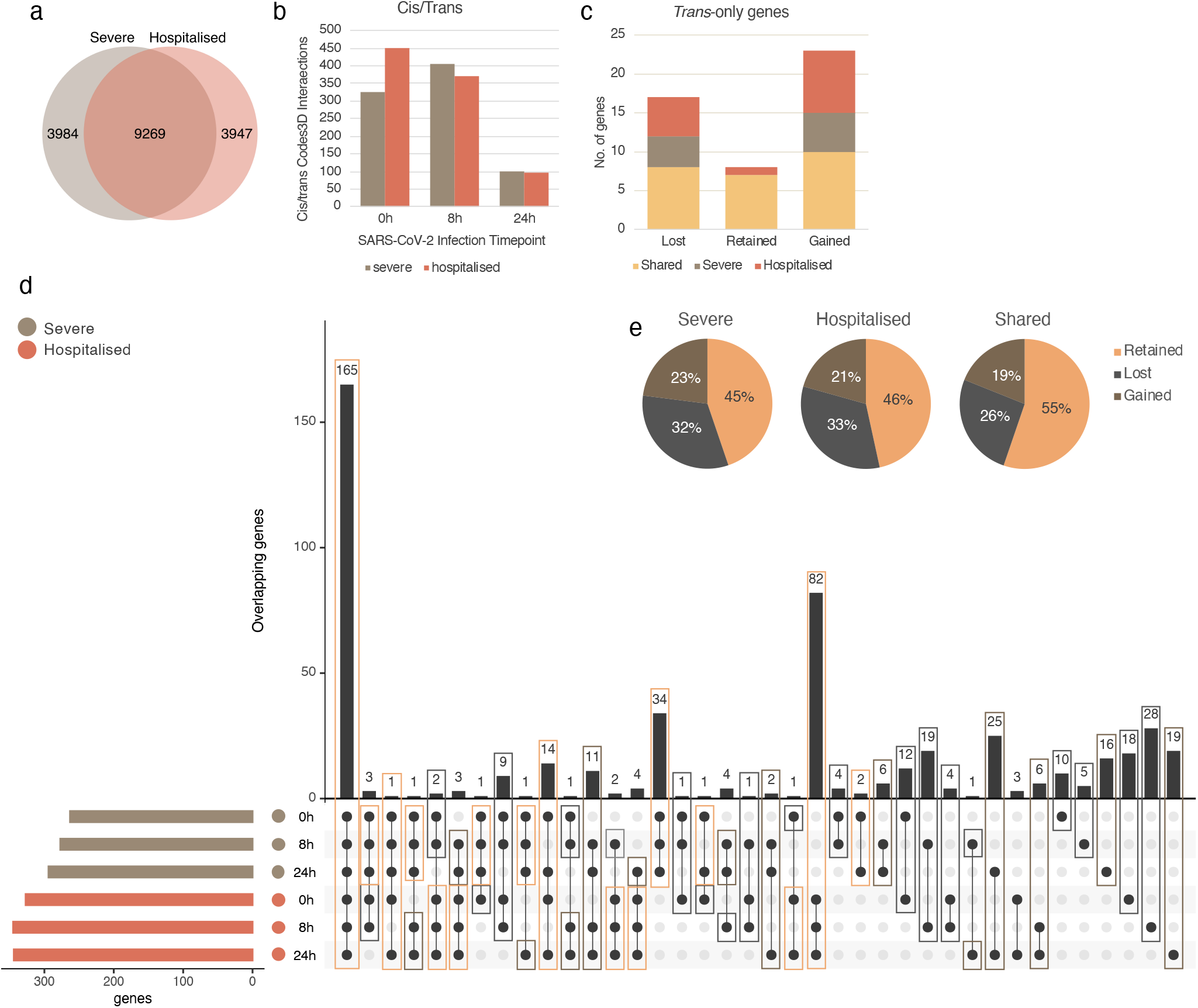
**(a)** Severe (69.9%) and hospitalised (70.1%) GWAS variants, along with those in LD (R^2^ > 0.8), overlap. **(b)** Ratio of cis/trans interactions across time. *Trans* SCeQTLs increase over time, consistent with weakening chromatin domains and deregulated loops. **(c)** Genes being targeted by *trans*-only SCeQTLs. 40 of 48 are either lost or gained across time. **(d)** The upset plot shows overlaps and distinct predicted gene targets. 165 shared, 34 severe and 82 hospitalised were retained following infection. Gene targets gained over time are highlighted in red. **(e)** Differential interactions (i.e. SNP-gene pairs) across time in severe (55%), hospitalised (54%), and shared (45%) groups.

**Supplementary figure 2.**
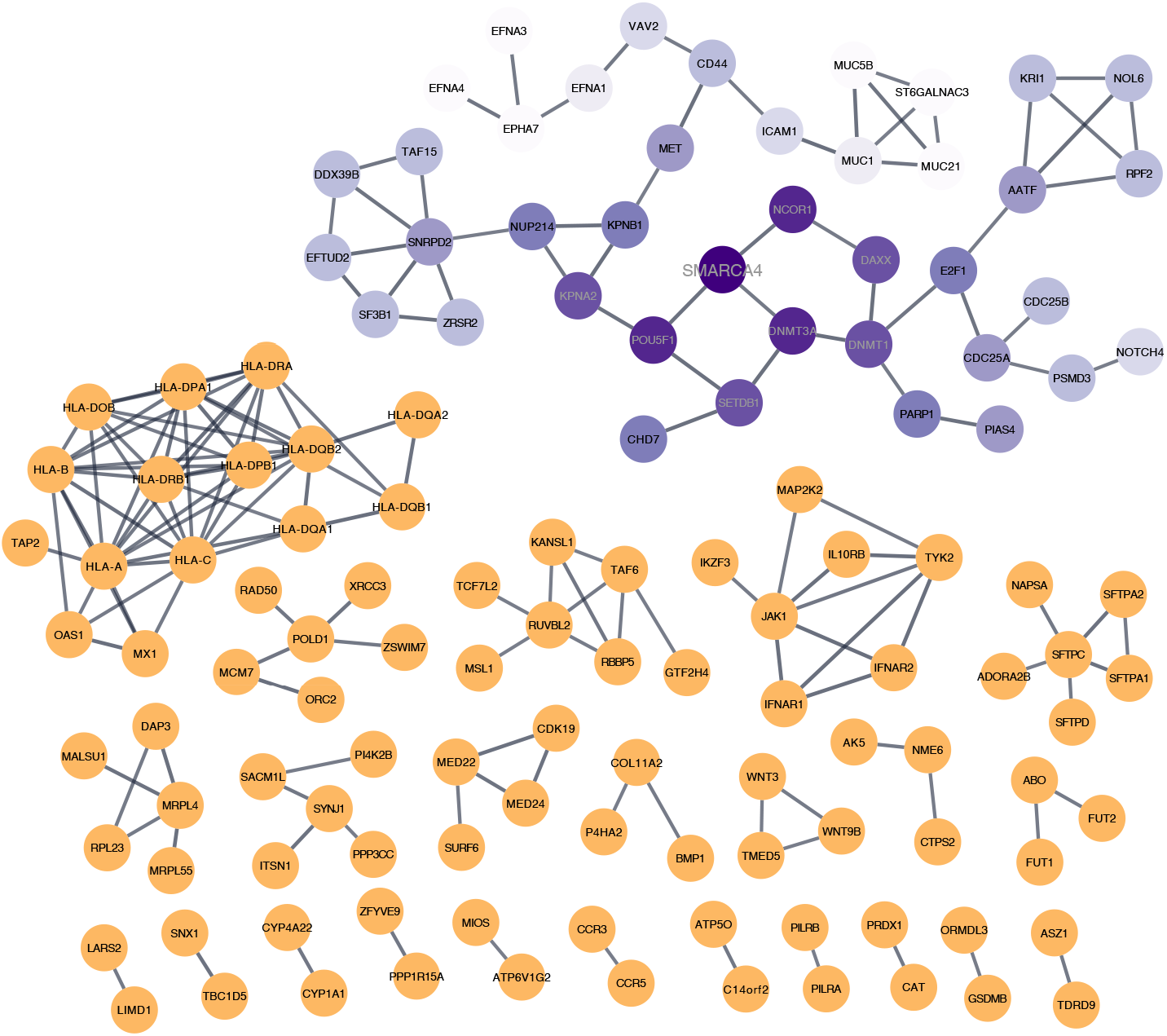
Protein-protein interaction network of all predicted target genes associated with germline risk of severe SARS-CoV-2 infection. The entire STRING^28^ database protein interaction network from severe and hospitalised genes expanded from Figure 3. Singletons were removed from the network. The largest within-network cluster is centered on SMARCA4 and its chromatin remodeller neighbours.

